# Developing internationally agreed core indicators for surveillance of preconception health: protocol for a consensus study

**DOI:** 10.64898/2026.01.27.26345014

**Authors:** Danielle Schoenaker, Jennifer Hall, Ghadir Fakhri Al-Jayyousi, Ana Luiza Vilela Borges, Palwende R. Boua, Jerry Kok Yen Chan, Tamar Chitashvili, Ilse Delbaere, Anastasia Demidova, Kassahun Fikadu, Sarah Fisher, Avishek Hazra, Sharon James, Chee Wai Ku, Eri Maeda, Zahid Memon, Daniel Munblit, Annemarie Mulders, Zivai Murira, Wendy V. Norman, Amy Ogle, Vani Sethi, Hessa Ibrahim Shahbic, Nadira Sultana Kakoly, Sarah Verbiest, Judith Stephenson, the International Core Indicators for Preconception Health and Equity (iCIPHE) Alliance

## Abstract

**Background:** Interventions and policies to optimise preconception health are increasing internationally. This stems from growing recognition that improving preconception health can improve maternal and child health outcomes and advance equity by reducing inequalities and address inequities for people of reproductive age and any children they may have. Interventions and policies should be evaluated through population-level surveillance of preconception health to inform the development of new initiatives, monitor effectiveness, and support advocacy for international adoption of successful strategies. However, there is currently no internationally agreed set of preconception health indicators available and suitable for international surveillance. The International Core Indicators for Preconception Health and Equity (iCIPHE) Alliance was established to address this gap.

**Aim:** To prioritise core indicators that can be used in low-, middle- and high-income countries for surveillance of preconception health and equity.

**Methods:** We held two workshops with the iCIPHE Alliance (multi-sectoral stakeholders from low-, middle- and high-income countries) to inform the design of this international consensus study. The development of core indicators will consist of three steps: (1) identifying an initial long-list of candidate surveillance indicators, and defining principles for scoring the importance of each indicator, through a literature review, public involvement, and workshops with iCIPHE Alliance members; (2) scoring each candidate indicator in terms of its importance for surveillance through a two-round Delphi survey among study participants; and (3) agreeing on the final core indicators through a series of consensus meetings with a selected group of study participants. We will recruit study participants from all World Health Organization (WHO) regions across four stakeholder groups: people of reproductive age (who do not belong to any of the other stakeholder groups); health and social care professionals; policy and programme professionals; and researchers.

**Ethical approval:** This study has been approved by the University of Southampton Faculty of Medicine Ethics Committee (ERGO 104447).

**Dissemination:** We will disseminate the priority core indicators through peer-reviewed publication, lay summaries, policy briefs and presentations. An implementation strategy, to enable monitoring of inequalities, inequities, and changes in preconception health over time within and between countries, will be developed with relevant national and international organisations to inform next steps.

## Introduction

Medical, behavioural and psychosocial factors before and between pregnancies (preconception health) can affect people’s own health and wellbeing, and their chance of successful pregnancy, pregnancy outcomes and the health and development of their children (1-5). Well-known preconception risk factors include, for example, lack of folic acid supplement use among women and anaemia, smoking, underweight and obesity, and teratogenic medication use among both women and men. These factors may increase risks of infertility, miscarriage, birth defects and preterm birth, as well as longer-term adverse cardiometabolic health outcomes for parents and children (3-5).

Maternal and paternal preconception risk factors are largely influenced by wider social, economic and environmental determinants, and inequalities in preconception health therefore need to be tackled through a dual approach to advance equity (6, 7). This includes public health policies that address structural barriers and promote equitable access to care and support for all people of reproductive age, such as mandatory folic acid fortification (8) as well as high quality and culturally safe health care to support individuals plan and prepare for pregnancy and parenthood (9, 10).

Advocacy and implementation of folic acid fortification, multiple micronutrient supplementation, and clinical preconception care guidelines, among other intervention strategies, are increasing internationally in response to the growing evidence of the health benefits for current and future generations (11, 12), and national and international calls for action (13-16). While further intervention development and more widespread implementation are needed in all countries, national and local programmes reduce inequalities and improve equities in preconception health are in place or emerging (8, 17-23).

The effectiveness of the growing number of programmes and policies being implemented to optimise preconception health globally should be monitored to provide evidence of impact (or lack thereof). Such evaluation is also needed to inform the development of and investment in new interventions, service delivery pathways and approaches to reach the most marginalised and ensure equitable benefit. This can be achieved through preconception health surveillance, which we define as: *“The ongoing systematic collection, analysis and interpretation of preconception health indicators at population and system levels to inform the planning, implementation and evaluation of efforts to optimise health before and between pregnancies, improve pregnancy outcomes and reduce health inequalities in (potential) parents and the next generation”*.

There are very few initiatives dedicated to surveillance of preconception health. Some specific individual indicators are reported through national and international surveillance platforms or annual reports, such as folic acid supplement use, obesity and anaemia (24-26). Selected countries, including England, The Netherlands and Qatar, are also developing surveillance systems that consolidate a comprehensive view of the population’s preconception health, care and support.

However, these individual systems may not include indicators that are relevant across low,- and middle- and high-income countries, that consider indicators relevant to equitable gain of preconception health and enable comparison. The first step towards a coordinated approach to surveillance of preconception health is the development of internationally agreed core preconception health indicators.

This paper describes the protocol for a study which aims to develop core indicators that can be used in low-, middle- and high-income countries for surveillance of preconception health and care, through an international consensus process among key stakeholder groups including people of reproductive age, health and social care professionals, policy professionals, and researchers.

## Methods

The International Core Indicators for Preconception Health and Equity (iCIPHE) Alliance was established in September 2022 (27). The iCIPHE Alliance has academic, clinical, policy/programme and public/community stakeholder representation from 24 low-, middle- and high-income countries (May 2025).

We held two workshops with Alliance members with representation from all six WHO regions (September 2022 [hybrid] and March 2023 [online]) to guide the aims, study design, stakeholder involvement and anticipated outcomes of this international consensus study (**Figure 1**).

**Figure 1.**
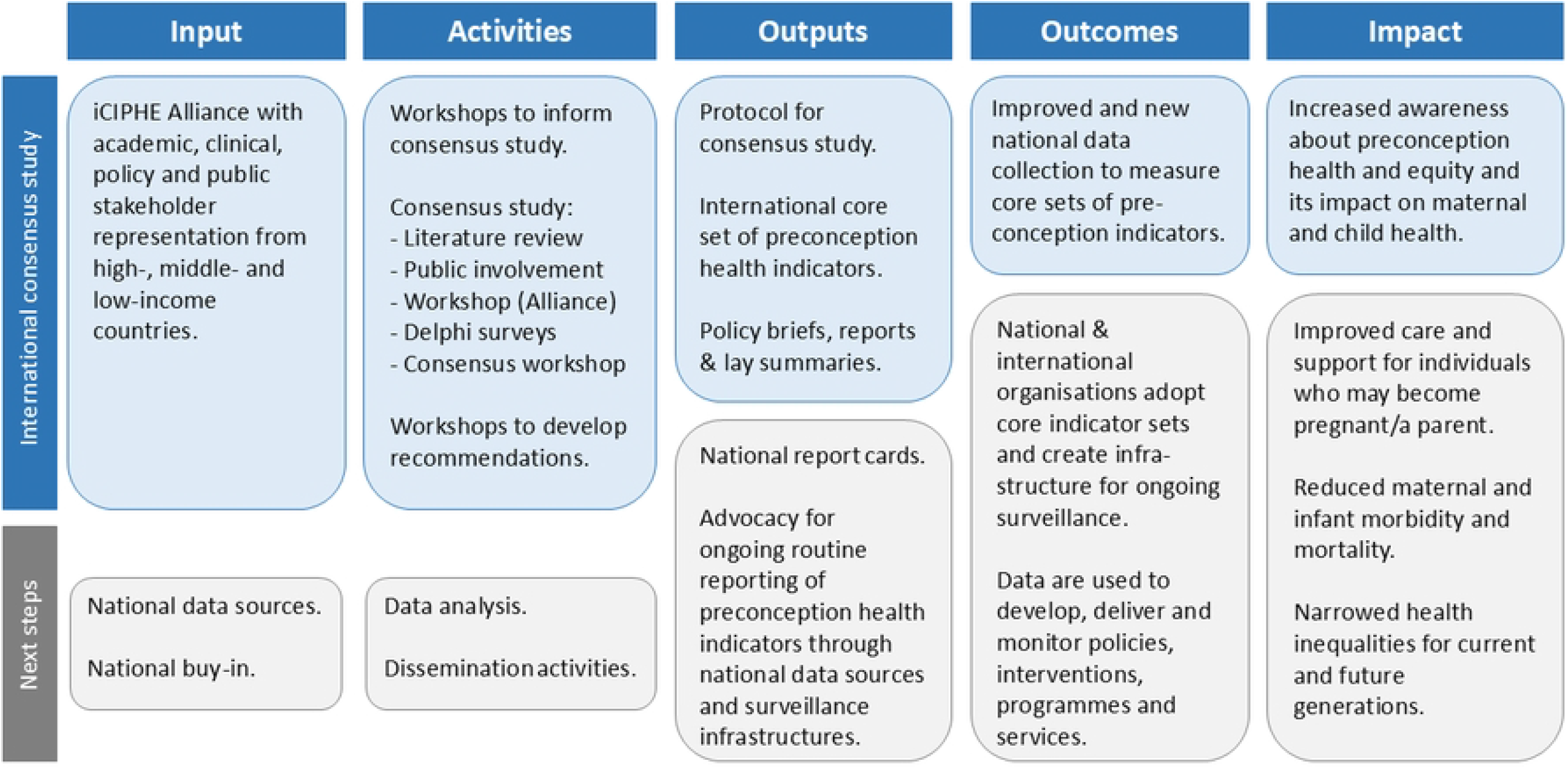
Logic model for an international consensus to develop core preconception health indicators, and next steps for the iCIPHE Alliance. iCIPHE, International Core Indicators for Preconception Health and Equity.

There is no standardised method for the development of core indicators. Where relevant, we used the Core Outcome Measures in Effectiveness Trials (COMET) handbook, Core Outcome Set-STAndards for Development (COS-STAD) and Core Outcome Set-STAndardised Protocol (COS-STAP) in the planning of the design of this study (28-30). Study findings will be reported according to Core Outcome Set-STAndards for Reporting (COS-STAR) recommendations (31). This protocol was prospectively registered with the COMET initiative (https://www.comet-initiative.org/Studies/Details/3725).

### Scope of the core indicator set

#### Population and timeframe

The core indicator set will include indicators of preconception health that are relevant to all people of reproductive age (female and male irrespective of gender identity, aged 15-49 years). Preconception health indicators can be defined as medical, behavioural and psychosocial risk factors or exposures, as well as broader determinants of health, that may affect potential future pregnancies among people of reproductive age irrespective of gender (32).

Depending on the indicator, the population may be sex-specific, and refined based on definitions proposed in the 2018 Lancet Series on preconception health: (1) weeks to months before conception (individual perspective – reflecting conscious intention to conceive) and (2) months or years before conception (public health perspective – reflecting longer periods needed to address preconception risk factors) (1). For example, preconception folic acid supplement use is recommended for women at least 3 months before trying to conceive, while diet and physical activity behaviours among prospective parents may need to be addressed earlier to ensure wider determinants (e.g. social and economic factors) and health and behaviours can be adequately addressed before conception. Where relevant, the population and timeframe will be included in the definition of example indicator measures.

Indicators may also include direct equity measures (e.g. indicators that allow preconception health measures to be compared across groups such as education and ethnicity) or system-level indicators that may reveal differences across groups (e.g. access to preconception healthcare).

#### Interventions

The core indicators will be relevant to interventions or actions that aim to improve and reduce inequalities in preconception health. The aim of preconception health and care interventions is to assist with pregnancy planning, improve pregnancy outcomes and the health and wellbeing of parents and the next generation (1, 13). Such interventions may be universal at the population-level (e.g. improving the food environment) and/or targeted at the individual-level (e.g. individualised preconception healthcare provided through primary care to patients intending to conceive) (6, 7, 10).

#### Context of use

The primary purpose of developing core indicators is population-level surveillance of preconception health to identify opportunities for preconception health gains, inform the development of new interventions, monitor effectiveness, and support advocacy for international adoption of successful strategies (Figure 1).

International consensus on core indicators will inform the development of national and international reports and surveillance infrastructures. Regular national snapshots of preconception health (including inequalities and/or inequities in preconception health), as well as cross-country comparison of core indicators and change over time, will also serve to hold governments, public health agencies and healthcare organisations to account for developing, delivering and evaluating policies, interventions, programmes and services that improve and reduce inequalities in the population’s preconception health.

Routine collection of health data varies widely across countries. Although many will already be routinely collected (at least in some countries), core indicators do not have to be limited to those currently measured and recorded in national data sources. We anticipate that the internationally agreed core preconception health indicators will support national advocacy for ongoing, improved and new routine data collection and reporting. The consensus process will be guided by scoring principles that do not rely on data availability. These scoring principles will be determined in Step 1 of the study (described below) and help guide study participants to determine the level of importance of each indicator for surveillance.

We will aim for the core indicators to be applicable to low-, middle- and high-income countries. We recommend that, where possible and pertinent, indicators are reported for the overall population of people of reproductive age, and for social, cultural and demographic subgroups to describe and monitor inequalities in preconception health. Given the variation in preconception health and care priorities across different countries, we anticipate the potential to identify a ‘*common core indicator set*’ of preconception health indicators relevant to all settings and recommended for global surveillance, as well as ‘*context-specific indicator sets*’ with additional indicators that may be considered for local surveillance.

#### Study overview

The international consensus study will consist of three steps: (1) identifying an initial long-list of candidate surveillance indicators, and defining principles for scoring the importance of each indicator, through a literature review, public involvement and workshops with iCIPHE Alliance members; (2) scoring each candidate indicator in terms of its importance for surveillance through a two-round Delphi survey among study participants; and (3) agreeing on the final core indicators through a series of consensus meetings with a selected group of study participants (**Figure 2**).

**Figure 2.**
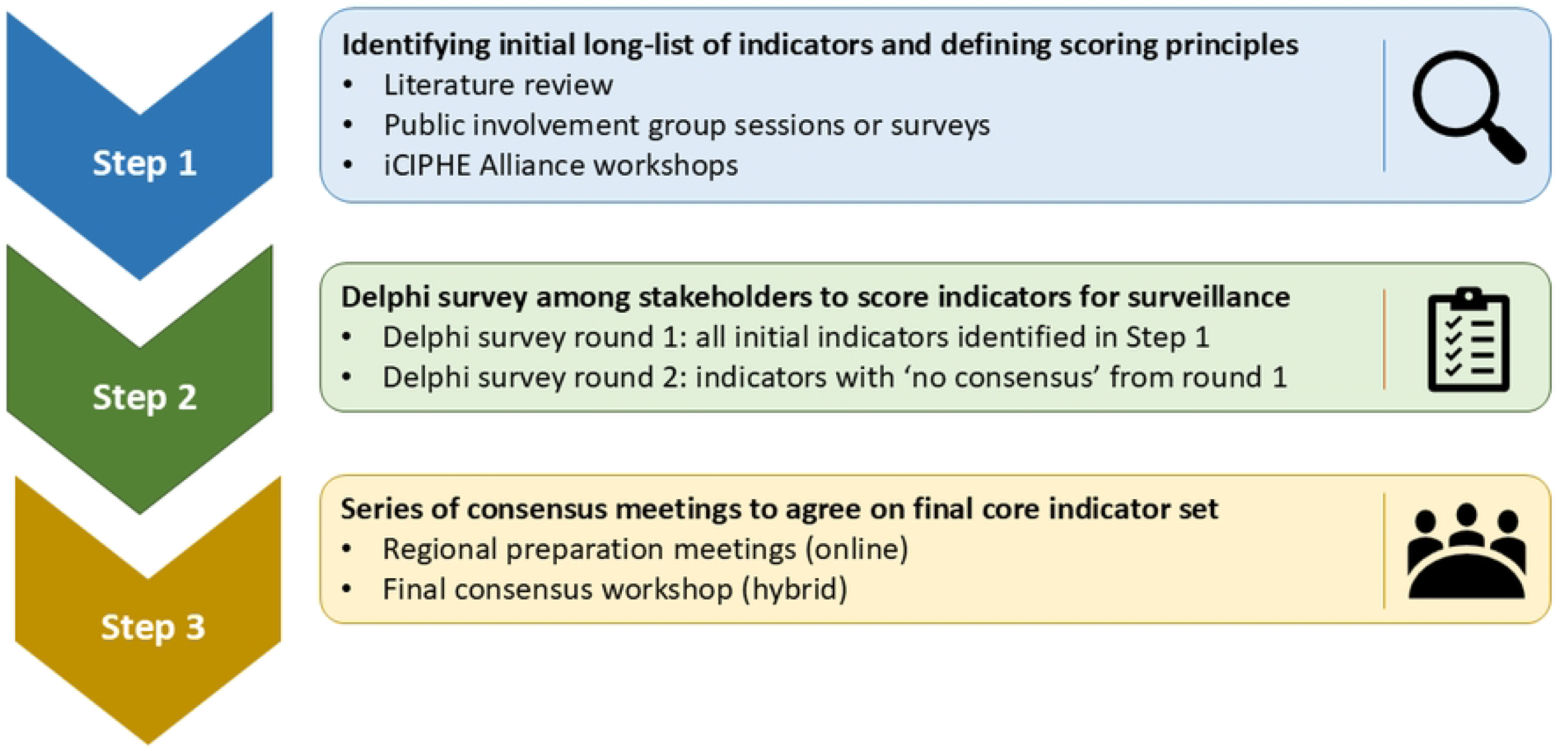
Flowchart of steps involved in development of core indicators. iCIPHE, International Core Indicators for Preconception Health and Equity.

### Step 1: Identify initial long-list of indicators and define scoring principles

#### Literature review

Given the recent publication of a review of preconception health indicators (32) and the extensive experience and knowledge in the field among iCIPHE Alliance members, a pragmatic approach will be used to identify the initial long-list of indicators and possible scoring principles based on published literature. All Alliance members will be asked to provide relevant publications for inclusion in the review, including reviews of preconception health indicators, preconception care guidelines, national preconception reports and consensus studies, and other relevant published documents.

In addition, we will conduct a scoping review to identify indicators reported in the preconception health and care literature from umbrella reviews and systematic reviews on maternal and paternal preconception health published in the past 5 years (Research Registry registration 1659) (33) and a systematic review to identify original studies that explore which preconception health indicators matter most to the public (PROSPERO registration CRD42023430759) (34).

Data extracted from relevant publications will produce a long-list of candidate preconception health indicators and examples of indicator measures, initially grouped across 12 topic areas/domains based on our recent review: wider determinants of health, health care, emotional and social health and support, reproductive health and family planning, health behaviours and weight, environmental exposures, cervical screening, immunisation and infections, mental health conditions, physical health conditions, medication use, and genetic risk (32). Data extraction will also inform an initial list of potential principles that may be used in the next step to score the importance of each indicator for surveillance.

#### Public involvement

Indicators identified in academic literature, policy documents and clinical guidelines represent indicators considered important to research, policy/programme and health professionals. Indicators or issues that matter most to the public (i.e. people of reproductive age in the general population whose preconception health we aim to optimise) may differ from published indicators, so public involvement of individuals with relevant lived experience will be pursued to ensure the initial list of indicators captures surveillance-appropriate indicators considered important to the public (28). Similarly, scoring principles identified through the literature (e.g. strength of evidence that indicator is associated with maternal and child health, or indicator is included in national policy or clinical guidelines) may not be relevant to the public. Discussions with members of the public (i.e. people of all genders of reproductive age irrespective of previous pregnancy experiences and future pregnancy and parenthood aspirations) will ensure that the guidance to complete the Delphi survey includes scoring principles relevant to their priorities and concerns.

We will obtain input from the public in each WHO region, through online or in-person group sessions, or through an anonymous online or paper-based survey, depending on local circumstances and capacity. Group sessions or distribution of surveys will be facilitated by a local iCIPHE Alliance member and coordinated by DS. We will aim to obtain input from 5-8 members of the public per WHO region. Convenience sampling will be used based on local access to public involvement groups by Alliance members, while aiming to obtain input from diverse individuals in terms of age, gender, ethnicity and health status.

Members of the public will be contacted by a local iCIPHE Alliance member, who will organise a group session or request survey completion. The group sessions will last for up to 1 hour or until no further new ideas are forthcoming. A topic guide and survey have been developed (Supplementary File 1). Both methods will identify preconception health indicators that are considered important to the public, as well as scoring principles based on how they decide what is most important to them. The topic guide and survey may be adapted and/or translated to suit local contexts. Members of the public will be reimbursed for their participation if appropriate and in line with local guidelines.

#### Workshops with iCIPHE Alliance

We will review the initial long-list of indicators, examples of indicator measures, and scoring principles generated from the literature review and public involvement with the iCIPHE Alliance during face-to-face and online workshops to agree on preconception health indicators to be included in the Delphi survey.

During these workshops, Alliance members may suggest additional indicators and examples of indicator measures to be added based on their local and international expert knowledge. Indicators may also be moved to a different or new domain, removed, combined to avoid redundancy, and wording and definitions refined for clarity, accuracy and relevance for the international context. If the long-list of indicators includes >50 items, then a ranking exercise will be used to prioritise up to 50 indicators for inclusion in the Delphi survey. The Core Working Group (see Acknowledgements) will make final decisions on the grouping and wording of the domains, indicators and examples of indicator measures. The list of scoring principles will also be discussed and potentially refined with Alliance members during the workshops. We will use a ranking exercise to define a maximum of five scoring principles to present to participants in the later Delphi survey for their consideration when scoring the importance of each indicator for surveillance. We will record these workshops to ensure accurate capture of all input.

### Step 2: Delphi survey

We will develop the core indicator set using a modified Delphi method. Delphi methodology is used to allow study participants with expert knowledge or lived experience on a particular subject to achieve convergence of opinion on the importance of different indicators using repeated survey rounds (28).

Members of the iCIPHE Alliance, including Core Working Group members, will be invited to participate in the Delphi survey and consensus meetings.

#### Recruitment of study participants

We will invite individuals from four stakeholder groups across each WHO region to participate in the online Delphi survey: (1) members of the public; (2) health and social care professionals; (3) policy and programme professionals; and (4) researchers. Members of the public will be individuals of any gender aged 18-49 years who do not belong to any of the other stakeholder groups or representative of the public for example through a non-profit or community-based organisation. The health and social care professionals group may include anyone caring for patients of reproductive age, such as primary care practitioners, obstetricians, midwifes, health visitors, sexual and reproductive health doctors, nurses, psychologists, nutritionists and social workers, as well as commissioners (i.e. professionals who prioritise, resource and monitor local and national health and social care services). Policy and programme professionals may work in government, public health agencies, healthcare organisations and charitable organisations, and be involved in, for example, national data collection and surveillance, health promotion, prevention, and advocacy. Researchers with expertise in relevant fields, such as preconception health, sexual- and reproductive health, public health, primary care and routine healthcare or population-based survey data, will also be invited to participate. Individuals will be asked to respond from their lived experience (group 1) or professional (group 2-4) perspectives. Participants’ stakeholder group, WHO region, and other characteristics (described below) will be identified through the Delphi survey. If a stakeholder group, WHO region or participant characteristic is poorly represented, more targeted recruitment may be undertaken (for example to ensure contributions from underserved groups such as Indigenous peoples), however recruitment will not be based on a specific sampling frame. The survey will be translated into local languages where possible. Study participants will not be reimbursed for their participation.

We will recruit participants across all WHO regions through relevant networks of iCIPHE Alliance members. Potential recruitment channels are listed in **Supplementary Table 1**, and include patient organisations, charities, professional organisations and national preconception health research networks. Participants will be identified through for example email distribution lists, newsletters and websites. Further recruitment through snowballing will be encouraged.

#### Sample size

There is no generally accepted guidance on the optimal sample size for Delphi surveys. Sample size is not based on statistical power but is a pragmatic choice (28). We aim to recruit around 25 study participants for each stakeholder group per WHO region for the first Delphi survey, to account for a 20% drop-out rate between the first and second survey round. This is feasible based on a review of international consensus studies (35). Retaining participants between survey rounds will be maximised through personalised email invitations, including only ‘No consensus’ items based on the first round in the second survey round, and adopting a minimum time between rounds (35, 36).

#### Surveys

We will invite participants to complete two consecutive rounds of the Delphi survey via email, which will include a participant information sheet explaining the objectives of the core indicator set, the principles that may be considered when scoring the importance of indicators for surveillance, and an outline of the study process. Study data will be collected and managed using Qualtrics, a secure web application for building and managing online surveys. Informed consent will be obtained from all participants who agree to take part, alongside participants’ name, email address and characteristics (stakeholder group, sex, ethnicity/ethnicity, country of residence, age group).

The first survey will include the long-list of all candidate indicators identified in step 1 grouped by domain for ease of completion. Participants will be asked to score the importance of each indicator for the purpose of surveillance using a 3-point scale: ‘No or limited importance’, ‘Important but not critical’ and ‘Critically important’. An ‘Unable to score/not applicable’ option will be included for each item for participants who are unclear or don’t have the knowledge to score specific indicators. Participants will also be given the opportunity to add a maximum of two additional indicators. A new indicator will be incorporated into the second survey round if it is suggested by two or more participants and considered distinct from already included items based on expert opinion of the Core Working Group.

After the first survey round, data will be analysed using descriptive statistics. Responses for each indicator will be summarised in simple bar graphs for each stakeholder group and fed back anonymously when participants are invited to complete the second survey round. During completion of the second survey, participants will be asked to reflect on the summary scores for all stakeholder groups and their own score, before deciding if they would like to keep or re-score each item. Only indicators that reached no consensus in the first round based on pre-specified definitions (see below) will be included in the second survey.

If a participant does not complete the second survey, their scores from the first survey will be retained in the study. The proportion of missing responses will be reported, and the impact of attrition bias will be assessed by comparing scores from participants who completed both rounds with those who only completed the first round.

#### Consensus definition

There are numerous ways to define consensus criteria (28). For our study, it is important that the majority of participants across most stakeholder groups agree that an indicator is of critical importance and only a small minority consider it to have limited importance for the indicator to be included in the core indicator set. The following consensus definition will therefore be applied to identify core indicators:

1. ‘Consensus in’ (classified as core indicator): ≥70% of participants in at least three stakeholder groups score the indicator ‘Critically important’ and ≤15% of participants in at least three stakeholder groups score the indicator ‘No or limited importance’
2. ‘Consensus out’ (do not classify as core indicator): ≥70% of participants in at least three stakeholder groups score the indicator ‘No or limited importance’ and ≤15% of participants in at least three stakeholder groups score the indicator ‘Critically important’
3. ‘No consensus’ (for further discussion in final consensus meeting): anything else

While we are not aiming for a pre-defined number of core indicators, the Core Working Group will review the consensus definition after the first survey round if they consider the number of indicators scored as ‘Consensus in’ to be too small or too large. Consensus definitions, such as the percentage cut-off of participants who need to score the indicator ‘Critically important’ or the number of stakeholder groups who score the indicators as ‘Critically important’ or ‘No or limited importance’, may be adjusted.

While we are aiming to reach global consensus, the consensus definition does not require agreement on core indicators across WHO regions as we anticipate this will be challenging. Differences in scoring of indicators among participants across WHO regions will be further discussed in regional preparation meetings and the final consensus workshop and addressed as part of a ‘*common core indicator set*’ and ‘*context-specific indicator sets*’.

#### Pilot survey

UK-based public contributors (Sarah F [co-author] and Tanjida R) will help shape the study materials through feedback, including on the participant information sheet, recruitment materials, wording of the survey questions, items and response scale format, and consensus meeting topic guide. To ensure ease of completion, acceptability and usability among international study participants, language-specific draft surveys will be completed by two Alliance members in each WHO region, who will ask two independent stakeholders (i.e. non-Alliance member and at least one member of the public) in their country/region to complete the survey and provide feedback. The feedback will be addressed in producing the final Delphi survey and study material.

### Step 3: Consensus meetings

We will hold a series of online consensus meetings to discuss the results of the Delphi survey and reach international consensus on a final core indicator set. At the end of the second Delphi survey, all English-speaking participants (not limited to iCIPHE Alliance members) will be asked if they are willing to take part in the consensus meetings. We will aim to have participant representation from each WHO region and stakeholder group, with purposive sampling if a large number of participants are interested in attending. Participants will be invited to the consensus meetings after the analysis of the second survey round has been completed. They will be informed of the logistics of attending, including partial or full reimbursement of expenses.

#### Regional preparation meetings

Preparation meetings may take place if different indicators are scored ‘consensus in’ and ‘consensus out’ across regions based on round 2 Delphi survey results (e.g. differences across WHO region, or high-income and low-and middle-income countries). There will be no specific criteria on how large these differences across regions need to be, but this will be discussed among Core Working Group members who will decide if regional preparation meetings are required. If this is the case, 2-hour regional online preparation meetings will be held to facilitate discussion and sharing of region-specific views ahead of the final consensus meeting. These meetings will be chaired by iCIPHE Alliance Core Working Group members and will not be recorded.

#### Final consensus workshop

The final 3-day hybrid consensus workshop will bring participants from all WHO regions and stakeholder groups together to discuss and vote on the final ‘*common core indicator set*’, and potential ‘*context-specific indicator sets*’ if needed based on Delphi survey results. We will aim to have one study participant from each of the four stakeholder groups across all six WHO regions (24-30 meeting participants). The workshop will be chaired by a non-voting independent experienced facilitator (i.e. someone outside the Alliance) and audio-recorded and transcribed using Zoom to contextualise decision making towards the final core indicators. Workshop participants may not be reimbursed for their participation.

Summary scores based on the Delphi survey will be shared with participants before the workshop and discussed during the workshop using nominal group technique (28). Participants will be randomly allocated into small groups to discuss the indicators. All participants are then brought together, and each participant will be asked to express their opinion on indicators considered for inclusion in the final core indicator set. This will be followed by voting using Mentimeter (37). Similar processes will be used to agree on the ‘*common core indicator set*’ and potential ‘*context-specific indicator sets*’, possibly with voting by WHO region.

### Ethical approval

This study has been approved by the University of Southampton Faculty of Medicine Ethics Committee (ERGO 104447). At the time of manuscript submission, ethical approval for the full study was granted and pilot testing of the survey was underway.

### Status and timeline of the study

A tentative timeline for the study is outlined in **Table 1**.

**Table 1.** Tentative timeline of the international consensus study*.

|  |  |
| --- | --- |
| April-July 2023 | Step 1: literature review |
| August 2023-April 2024 | Step 1: public involvement to inform the Delphi survey items |
| September-October 2024 | Step 1: workshops with iCIPHE Alliance |
| November 2024-September 2025 | Obtaining ethics approvals, including survey translations |
| September-December 2025 | Step 2: pilot survey |
| February-March 2026 | Step 2: recruitment & Delphi survey round 1 |
| April 2026 | Step 2: analysis of Delphi survey round 1 |
| May-June 2026 | Step 2: Delphi survey round 2 |
| July 2026 | Step 2: analysis of Delphi survey round 2 |
| September-October 2026 | Step 3: consensus preparation meetings |
| November 2026 | Step 3: final consensus workshop |
| December 2026 onwards | Dissemination and implementation |

\* Public contributors will be involved in all steps of the study, including Delphi survey and study participant material development (Step 1), pilot testing of the Delphi survey, study participant recruitment, and as study participants (Step 2), preparation of materials for the consensus meetings and workshop and as workshop participants (step 3), and to inform, design and share study findings and the implementation plan.

## Discussion

### Strengths

There is currently no internationally recognised set of preconception health indicators that could be used for surveillance. Informed by input from the iCIPHE Alliance, our study will be the first to combine insights from multiple stakeholder groups across low-, middle- and high-income countries to develop consensus on core indicators that can be used globally.

Priority preconception health indicators have been identified in the UK, USA and Netherlands based on input from researchers, health professionals and policy professionals, however, members of the public (who do not belong to any of the other stakeholder groups) were not consulted in these exercises. Our study will involve members of the public across all WHO regions throughout the study including before (e.g. feedback on the pilot survey), during (e.g. participation in all three steps of the consensus study), and after (e.g. dissemination of findings through lay summaries). This approach strives to provide opportunities for the public (including marginalized groups usually excluded from indicator development) to contribute to and inform consensus on the development of core indicators.

Previous studies have considered data availability (UK and USA) and data completeness (UK) in the selection of core preconception health indicators. Based on discussion with the iCIPHE Alliance, the development of our core indicators will initially not rely on the availability of relevant data as national data on preconception health are not currently collected in many countries. An important aim of the internationally agreed core indicators is to support advocacy for improved and/or new standardised national data collection to measure preconception health indicators and thereby facilitate the development of surveillance infrastructures across low-, middle- and high-income countries.

### Limitations and potential for mitigation

Preconception health covers a wide range of medical, behavioural and psychosocial factors and wider determinants, and some of these may be country-specific, such as cultural practices, HIV status and malaria. Moreover, health and policy priorities differ across countries, and in some cases across federations, which may influence the perceived importance of preconception health indicators for national surveillance. This may challenge our ability to reach international consensus on a core indicator set. If ‘No consensus’ is reached for most indicators after the first Delphi survey round, the Core Working Group will re-consider and potentially adapt the consensus definition. Survey data will be analysed by region to inform discussions during the regional preparation meetings and final consensus workshop. We anticipate that not all indicators included in the final ‘common core indicator set’ will be applicable to all countries. Equity considerations will require that countries or regions have flexibility to develop ‘context-specific indicator sets’ that respect local population needs, capacities and structural conditions. Further country or regional prioritisation may still be needed to consider local health and health system issues, cultural factors and government priorities.

We will aim to achieve diverse representation of study participants throughout the study by using multiple recruitment sources and active engagement with underserved community groups. For practical reasons, the survey will be available in a limited number of languages (depending on funding and/or local translations by iCIPHE Alliance members) and the consensus meetings will be held in English only which may disproportionately affect participation of linguistically marginalised groups. We will document these constraints as limitations of our final agreed core indicator set. Participants will be adults of reproductive age, and those aged <18 will not be able to take part. Moreover, the use of online platforms may lead to underrepresentation of digitally disadvantaged groups and regions with limited access to digital resources. The global nature of the study will challenge participation from international participants across the various time zones in the final consensus workshop, however, participants will be made aware of this when asked about participation, and regional preparation meetings will facilitate sharing and documenting of region-specific views ahead of the final consensus workshop.

### Dissemination plan and next steps

The results of the literature review, public involvement, workshops with the iCIPHE Alliance, Delphi survey and consensus meetings will be published in peer-reviewed open access journals. These will be circulated, alongside translated lay summaries, to all study participants and members of the iCIPHE Alliance for local dissemination. Findings will also be presented at national and international conferences and meetings.

We are aiming for the international core set of preconception health indicators to be used in low-, middle- and high-income countries to produce national reports, and to be adopted by international organisations such as WHO. A dissemination and implementation strategy will be developed with the iCIPHE Alliance and relevant national and international organisations. A range of methods may be used, including for example workshops to develop international and national recommendations on data collection and ongoing monitoring of core indicators, with specific attention to enabling low-resource settings to adopt and operationalise the core indicators. Policy briefs and reports may be produced to raise awareness of core indicators and promote adoption. All dissemination and promotion material will be available on the iCIPHE Alliance webpage.

Following dissemination, an approach will need to be developed to monitor uptake of the core indicators in national data sources and surveillance infrastructures. This will include assessing whether inequities in data system capacity limit adoption, and where additional support or adaptation is needed. Over time, regular reviews of the core indicators will be needed to ensure they are up-to-date with the latest scientific evidence and clinical practice and respond to population changes and emerging health priorities.

## Conclusion

Through international multi-disciplinary collaboration and commitment to improving preconception health and equity, the iCIPHE Alliance has developed the first protocol for a consensus study to reach agreement on a core set of preconception health indicators for surveillance. This study will build on past and ongoing surveillance efforts and experiences in the UK, USA, Netherlands and Australia, and responds to calls for preconception health surveillance in other countries. We will use comprehensive methodology in line with recommendations for development of core outcome sets (28-30), and involve multiple stakeholder groups and end users from the start through to dissemination of the study findings. The proposed core indicators will serve to support advocacy for standardised national routine data collection of preconception health indicators and thereby enable national and international surveillance that can inform development of preconception health services and identify and address inequalities and inequities. Their monitoring over time within and between countries will provide an opportunity to inform the development and evaluation of interventions and ultimately contribute to reducing and narrowing inequalities in maternal and infant morbidity and mortality globally.

## Data Availability

N/A

## Acknowledgements

The authors would like to thank iCIPHE Alliance members who contributed to the idea and design of this consensus study through the workshops. We would also like to thank the University of Southampton Public Advisory Group of people of reproductive age who contributed to improvements in the public relevance and understanding of the study materials.

iCIPHE Alliance Core Working Group members include: Danielle Schoenaker, Judith Stephenson (co-chairs); Palwende R. Boua, Kassahun Fikadu (African Region); Wendy V. Norman, Amy Ogle, Sarah Verbiest, Tamar Chitashvili (Region of the Americas); Nadira Sultana Kakoly, Vani Sethi, Zivai Murira (South-East Asia Region); Jennifer Hall, Annemarie Mulders (European Region); Ghadir Fakhri Al-Jayyousi, Zahid Memon (Eastern Mediterranean Region); Sharon James, Chee Wai Ku, Eri Maeda (Western Pacific Region).

## Author contributions

DS and JS led the design of this study, and DS led the writing of the first draft of the paper. All co-authors contributed to informing critical aspects of the study protocol and to review and editing of the paper for important intellectual content.

## References

1. Stephenson J, Heslehurst N, Hall J, Schoenaker DA, Hutchinson J, Cade JE, et al. Before the beginning: nutrition and lifestyle in the preconception period and its importance for future health. Lancet. 2018;391(10132):1830–41.

2. Fleming TP, Watkins AJ, Velazquez MA, Mathers JC, Prentice AM, Stephenson J, et al. Origins of lifetime health around the time of conception: causes and consequences. Lancet. 2018;391(10132):1842–52.

3. Daly M, Kipping RR, Tinner LE, Sanders J, White JW. Preconception exposures and adverse pregnancy, birth and postpartum outcomes: Umbrella review of systematic reviews. Paediatr Perinat Epidemiol. 2022;36(2):288–99.

4. Carter T, Schoenaker DA, Adams J, Steel A. Paternal preconception modifiable risk factors for adverse pregnancy and offspring outcomes: a review of contemporary evidence from observational studies. BMC Public Health. 2023;23(1):509.

5. Caut C, Schoenaker D, McIntyre E, Vilcins D, Gavine A, Steel A. Relationships between Women’s and Men’s Modifiable Preconception Risks and Health Behaviors and Maternal and Offspring Health Outcomes: An Umbrella Review. Semin Reprod Med. 2022;40(3-04):170–83.

6. Stephenson J, Vogel C, Hall J, Hutchinson J, Mann S, Duncan H, et al. Preconception health in England: a proposal for annual reporting with core metrics. Lancet. 2019;393(10187):2262–71.

7. Barker M, Dombrowski SU, Colbourn T, Fall CHD, Kriznik NM, Lawrence WT, et al. Intervention strategies to improve nutrition and health behaviours before conception. Lancet. 2018;391(10132):1853–64.

8. Wald NJ. Folic acid and neural tube defects: Discovery, debate and the need for policy change. J Med Screen. 2022;29(3):138–46.

9. Dorney E, Boyle JA, Walker R, Hammarberg K, Musgrave L, Schoenaker DA, et al. A Systematic Review of Clinical Guidelines for Preconception Care. Semin Reprod Med. 2022;40(3-04):157–69.

10. Withanage NN, Botfield JR, Srinivasan S, Black KI, Mazza D. Effectiveness of preconception interventions in primary care: a systematic review. Br J Gen Pract. 2022;72(725):e865–e72.

11. Norris S, Conti G, Kumaran K, Barker M, Chou D, Craig A, et al. Optimising preconception health and care: strategies to improve lifelong health. In preparation.

12. Hazra A, et al. Policies and programmes to improve preconception nutrition in South Asia. Lancet Reg Health Southeast Asia. 2025.

13. World Health Organization. Preconception care: Maximizing the gains for maternal and child health. Policy brief. 2013. Available from: https://www.who.int/publications/i/item/WHO-FWC-MCA-13.02. [Accessed 23/06/2023].

14. World Health Organization. Global Strategy for Women’s, Children’s and Adolescent’s Health, 2016–2030, 2015. New York: United Nations. Available from https://www.who.int/life-course/partners/globalstrategy/en/. [Accessed 23/06/2023].

15. United Nations. Transforming our World: The 2030 Agenda for Sustainable Development, 2015. New York: United Nations. Available from https://sustainabledevelopment.un.org/post2015/transformingourworld/publication. [Accessed 23/06/2023].

16. Public Health England. Making the case for preconception care. 2018. Available from: https://assets.publishing.service.gov.uk/government/uploads/system/uploads/attachment_data/file/729018/Making_the_case_for_preconception_care.pdf. [Accessed 23/06/2023].

17. World Health Organization. Meeting to develop a global consensus on preconception care to reduce maternal and childhood mortality and morbidity. 2012. Available from: https://apps.who.int/iris/handle/10665/78067. [Accessed 23/06/2023].

18. Walani SR, Moley KH. Global strategies for change. In Preconception Health and Care: A Life Course Approach (eds. Shawe J, Steegers E, Verbiest S), 2020; pp. 287–297. Springer Nature Switzerland.

19. Ebrahim SH, Lo SS, Zhuo J, Han JY, Delvoye P, Zhu L. Models of preconception care implementation in selected countries. Matern Child Health J. 2006;10(5 Suppl):S37–42.

20. Ministry of Health, Welfare and Sport. ‘Solid Start’ action programme (Actieprogramme Kansrijke Start). Available from: https://www.kansrijkestartnl.nl/actieprogramma-kansrijke-start. [Accessed 18/08/2024].

21. Hall J, Chawla M, Watson D, Jacob CM, Schoenaker D, Connolly A, et al. Addressing reproductive health needs across the life course: an integrated, community-based model combining contraception and preconception care. Lancet Public Health. 2023;8(1):e76–e84.

22. World Health Organization (WHO). Reframing care and services to improve preconception health: meeting report, Geneva, Switzerland, 8-9 May 2024. Geneva: World Health Organization; 2024. Available from: https://www.who.int/publications/i/item/9789240101036. [Accessed 18/08/2024].

23. UNICEF South Asia. Advancing Preconception Nutrition in South Asia: Technical Brief. Kathmandu: UNICEF South Asia; 2025. Available from: https://www.unicef.org/rosa/reports/advancing-preconception-nutrition-south-asia-technical-brief. [Accessed 12/05/2025].

24. World Health Organization (WHO). Global Health Observatory. Available from https://www.who.int/data/gho/data/themes/topics/indicator-groups. [Accessed 18/08/2024].

25. UK Government Office for Health Improvement and Disparities. Fingertips - Public Health Data. Child and Maternal Health profile. Available from https://fingertips.phe.org.uk/child-health-profiles. [Accessed 18/08/2024].

26. Australian Institute of Health and Welfare. Mothers and babies. Available from https://www.aihw.gov.au/reports-data/population-groups/mothers-babies/overview. [Accessed 18/08/2024].

27. iCIPHE Alliance. Available from: https://www.ukpreconceptionpartnership.co.uk/about-us/iciphe-alliance. [Accessed 23/06/2023].

28. Williamson PR, Altman DG, Bagley H, Barnes KL, Blazeby JM, Brookes ST, et al. The COMET Handbook: version 1.0. Trials. 2017;18(Suppl 3):280.

29. Kirkham JJ, Davis K, Altman DG, Blazeby JM, Clarke M, Tunis S, et al. Core Outcome Set-STAndards for Development: The COS-STAD recommendations. PLoS Med. 2017;14(11):e1002447.

30. Kirkham JJ, Gorst S, Altman DG, Blazeby JM, Clarke M, Tunis S, et al. Core Outcome Set-STAndardised Protocol Items: the COS-STAP Statement. Trials. 2019;20(1):116.

31. Kirkham JJ, Gorst S, Altman DG, Blazeby JM, Clarke M, Devane D, et al. Core Outcome Set-STAndards for Reporting: The COS-STAR Statement. PLoS Med. 2016;13(10):e1002148.

32. Schoenaker DA, Stephenson J, Connolly A, Shillaker S, Fishburn S, Barker M, et al. Characterising and monitoring preconception health in England: a review of national population-level indicators and core data sources. J Dev Orig Health Dis. 2022;13(2):137–50.

33. Pitafi S, Schoenaker D, Hall J, Stewart C, Stephenson J. Umbrella Review of Global Preconception Care Indicators and Guidelines. Research Registry 2023 #reviewregistry1659. Available from: https://www.researchregistry.com/browse-the-registry#registryofsystematicreviewsmeta-analyses/registryofsystematicreviewsmeta-analysesdetails/649adfd25e1998001dd7c867/. [Accessed 29/06/2023].

34. Pitafi S, Schoenaker D, Hall J, Stewart C, Stephenson J. Determining which preconception health indicators are important to people of reproductive age. PROSPERO 2023 CRD42023430759. Available from: https://www.crd.york.ac.uk/prospero/display_record.php?ID=CRD42023430759. [Accessed 23/06/2023].

35. Gargon E, Crew R, Burnside G, Williamson PR. Higher number of items associated with significantly lower response rates in COS Delphi surveys. J Clin Epidemiol. 2019;108:110–20.

36. Hall DA, Smith H, Heffernan E, Fackrell K. Recruiting and retaining participants in e-Delphi surveys for core outcome set development: Evaluating the COMiT’ID study. PLoS One. 2018;13(7):e0201378.

37. Mentimeter. Available from: https://www.mentimeter.com/. [Accessed 23/06/2023].

